# Identification of A Novel CG307 Sub-clade in Third Generation Cephalosporin Resistant *Klebsiella pneumoniae* Causing Invasive Infections in the United States

**DOI:** 10.1101/2023.11.22.23298833

**Authors:** Selvalakshmi Selvaraj Anand, Chin-Ting Wu, Jordan Bremer, Micah Bhatti, Todd J Treangen, Awdhesh Kalia, Samuel A Shelburne, William C Shropshire

## Abstract

Despite the notable clinical impact, recent molecular epidemiology regarding third-generation cephalosporin-resistant *Klebsiella pneumoniae* (3GC-R*Kp*) in the United States remains limited. We performed whole genome sequencing of 3GC-R*Kp* bacteremia isolates collected from March 2016 to May 2022 at a tertiary care cancer center in Houston, TX using Illumina and Oxford Nanopore Technologies platforms. A comprehensive comparative genomic analysis was performed to dissect population structure, transmission dynamics, and pan-genomic signatures of our 3GC-R*Kp* population. Of the 194 3GC-R*Kp* bacteremias that occurred during our study timeframe, we were able to analyze 153 (79%) bacteremia isolates, 126 initial and 27 recurrent isolates respectively. While isolates belonging to the widely prevalent clonal group (CG) 258 were rarely observed, the predominant clonal group, CG307, accounted for 37 (29%) index isolates and displayed a significant correlation (Pearson correlation test *P*-value = 0.03) with the annual frequency of 3GC-R*Kp* bacteremia. Within our CG307 cohort, 89% (33/37) of our isolates belong to the global rather than previously described Texas-specific clade. Strikingly, we identified a new CG307 sub-clade (*i.e.,* cluster 1 isolates) comprised of 18 isolates characterized by the chromosomally-encoded *bla*_SHV-205_ and unique accessory genome content. This CG307 sub-clade was detected in various United States regions, with genome sequences from 24 additional strains becoming recently available in the NCBI SRA database. Collectively, this study underscores the emergence and dissemination of a distinct CG307 sub-clade that is a prevalent cause of 3GC-R*Kp* bacteremia among cancer patients seen in Houston, TX and has recently been isolated throughout the United States.

**DATA SUMMARY:** WGS data sequenced during this study period was submitted to NCBI and can be accessed within BioProject PRJNA648389. WGS data from previous study of carbapenem non-susceptible *Enterobacterales* can be accessed from BioProject PRJNA836696. Assembly information and BioAccession numbers are provided in Table S1.

**IMPACT STATEMENT:** Infections due to 3^rd^ generation cephalosporin resistant *Klebsiella pneumoniae* (3GC-R*Kp*) are considered among the most urgent public health threats. However, molecular epidemiology studies on 3GC-R*Kp* in the United States are limited. Our analysis indicates a preponderance of genetically diverse 3GC-R*Kp* isolates harboring the key antimicrobial resistance determinant *bla*_CTX-M-15_ at our institution. Importantly, however, we detected evidence of long duration transmission of highly genetically related CG307 and CG29 specific clusters at our institution. Interestingly, we rarely detected the pandemic CG258 lineage in our cohort and did not detect more than two genetically related CG258 isolates from this lineage. We found that 90% of our isolates from the most prevalent clonal group, CG307, belonged to a novel, nested-population of a “global” CG307 clade in contrast to the more commonly detected “Texas-specific” clade that has circulated in our region. We searched the NCBI SRA database using genomic markers of the novel CG307 clade and found evidence of this clade causing recent invasive infections in other locations across the United States. Our study highlights the shifting population dynamics of *K. pneumoniae* causing invasive infections and the necessity to continue AMR surveillance in order to identify emerging high-risk populations.

## INTRODUCTION

The necessity to address antimicrobial resistant (AMR) infections as a global public health threat has become increasingly apparent during the 21^st^ century. A recent study estimated that 4.95 million deaths were associated with AMR infections worldwide with *Klebsiella pneumoniae* infections being the third most common pathogen associated with mortality [1]. *K. pneumoniae*, a member of the *Enterobacterales* family, is an opportunistic pathogen that has the capacity to develop resistance to multiple classes of antibiotics. A recent meta-analysis estimated about 33% of nosocomial *K. pneumoniae* infections are caused by MDR strains highlighting the impact of these pathogens in the healthcare setting [2].

MDR *K. pneumoniae* clonal populations can acquire β-lactamase encoding genes such as extended-spectrum β-lactamase (ESBL) or AmpC encoding genes via horizontal gene transfer (HGT), which results in third-generation cephalosporin resistance (3GC-R). These HGT plasmid vectors can also harbor carbapenemase encoding genes such as the *bla*_KPC,_ which encodes the *Klebsiella pneumoniae* carbapenemase (KPC) that effectively hydrolyzes most classes of β-lactams [3-12]. One of the most concerning MDR *K. pneumoniae* clonal populations is the pandemic group of strains known as clonal group 258 (CG258), which include sequence types 258 (ST258), ST512, and ST11, that are all closely related (*i.e.,* the CG258 average pairwise SNP distance is ∼214) and strongly associated with worldwide *bla*_KPC_ dissemination [6, 13-17]. ST258 strains are the predominant MDR *K. pneumoniae* detected in the United States over the past two decades [5, 7, 16-19].

While ST258 prevalence has remained high in certain regions of the United States, there is growing evidence of an emergent CG307 lineage co-circulating with CG258 in the Houston, TX region [7, 9, 20, 21]. When performing comparative genomics between the two lineages, a discerning factor is the strong association of the ESBL encoding gene *bla*_CTX-M-15_ that is present in >90% of CG307 isolates whereas *bla*_CTX-M-15_ is detected in <10% of CG258 isolates [9, 10, 20]. Recent molecular epidemiological investigations of CG307 isolates have identified two predominant CG307 clades with a strong phylogenetic signal that distinguishes isolates of Texas origin from those from the more globally disseminated clade [10, 20]. When comparing the two ST307 lineages, the Texas-specific clade has a stable chromosomal insertion of two IS*Ecp1-bla*_CTX-M-15_ transposition units whereas the more globally disseminated CG307 lineage harbors *bla*_CTX-M-15_ primarily on large, multireplicon, F-type conjugative plasmids [8, 10, 20]. Furthermore, recent AMR surveillance studies from Europe have highlighted how CG307 strains are the primary drivers of 3GC-RKp (3GC-R*Kp*) infections within hospitals in part due to their strong association with *bla*_CTX-M-15_ carriage and transmission [22-24].

Given the high prevalence of CG307 in Houston and surrounding areas [7, 9, 20, 21, 25, 26] as well as a lack of current US-based 3GC-R*Kp* surveillance studies, we sought to use WGS to determine the molecular epidemiology of 3GC-R*Kp* causing bacteremia in cancer patients at our institution within a five-year timeframe. We found that CG307 strains were the most common CG detected whereas the previously highly prevalent CG258 was rarely identified. Interestingly, 90% of CG307 bacteremias were caused by strains of the global clade; in particular, we identified that a previously unidentified, global sub-clade with distinct genomic signatures caused approximately 50% of the CG307 bacteremias at our institution. Search of publicly available WGS data identified isolates of this sub-clade recently collected from geographically diverse sites throughout the United States. These data expand upon the rapidly changing epidemiology of 3GC-R*Kp*, including the increasing importance of emerging, diverse CG307 sub-clades.

## METHODS

### Study design and sample collection

The study included all K. *pneumoniae* bacteremias that occurred from March 1^st^, 2016, chosen coincident with the date of implementation of the Epic Electronic Health Record system at the University of Texas MD Anderson Cancer Center (MDACC) in Houston, TX, to May 30^th^, 2022. We defined an index *K. pneumoniae* bacteremia isolate as first occurrence of a positive blood culture and a recurrent *K. pneumoniae* bacteremia isolate as a positive blood culture that occurred at least 14 days from a previous *K. pneumoniae* bacteremia isolate. Antibiotic susceptibility testing of all *K. pneumoniae* bacteremia isolates was performed by the MDACC microbiology laboratory using the Accelerate PhenoTest™ BC (Accelerate Diagnostics), ETEST® (bioMérieux), and VITEK® 2 (bioMérieux) with susceptibility interpretations based on CLSI guidelines [27]. *K. pneumoniae* bacteremia isolates were considered as extended-spectrum cephalosporin resistant (3GC-R) if they had a ceftriaxone (CRO) minimum inhibitory concentration (MIC) ≥ 4 μg/mL and/or a predicted ESBL phenotype. Out of the 194 3GC-R*Kp* causing bacteremia during the study period, there were 79% (161/194) 3GC-R*Kp* isolates available for sequencing. Further information on sampling is included in the results section.

### Short-read and long-read sequencing

3GC-R*Kp* isolates were stocked in thioglycolate broth supplemented with 40% glycerol as part of an ongoing surveillance of bacteremia isolates. This surveillance study has received ethical approval through the University of Texas MD Anderson Cancer Center (MDACC) Institutional Review Board (Protocol ID: PA13-0334). MicroBank (MB) numbers are unique to patient infections and deidentified. Samples were plated onto Trypticase Soy Agar (TSA with Sheep Blood). These plates were subsequently incubated overnight at 37°C and 4% CO_2_. After 12-24 hours, a single colony was inoculated in autoclaved Miller’s LB Broth and incubated at 37°C for 2 hours with mild agitation (225 rpm). Two mL of the inoculated media were aliquoted into 15 mL Eppendorf tubes and spun down to pellet cells. Genomic DNA (gDNA) was extracted according to the protocol specified on QIAGEN “Blood and Cell Culture DNA Kit.” (Cat No. 69581).

ESC-R-*K. pneumoniae* isolates were sequenced by Illumina NovaSeq 6000 as previously described [21]. There were 126 3GC-R*Kp* isolates with 150 bp paired-end reads that passed quality control as assessed using fastqc-v0.11.9 (https://github.com/s-andrews/FastQC). We performed long-read sequencing on nine isolates of interest that were part of transmission clusters to obtain their complete genomes using the Oxford Nanopore Technologies (ONT) MinION platform as described previously [21]. Briefly, genomic DNA extracted for the purpose of short-read sequencing was used as input for ONT long-read sequencing. Library preparation was accomplished using the Rapid Sequencing Kit 96 V10 (SQK-RBK110.96). The input gDNA was normalized to 50 ng to ensure even distributions of libraries across pooled samples. The prepared libraries were loaded onto the R9.4.1 flowcell (FLO-MIN106D) for sequencing with MinKNOW software to generate fast5 files. Guppy-v6.4.6 basecaller was used to perform basecalling from the fast5 files to obtain fastq files using super high accuracy model (SUP).

### Short and long-read sequencing data analysis

Quality assessed, paired-end 150 bp reads were assembled via SPAdes-v3.15.5 [28] with default parameters and the inclusion of the isolate option. The quality assessment, assignment of sequence types, clonal groups, and capsule type of isolates was assessed using Kleborate-v2.2.0 with Kaptive-v2.0.0 activated [29, 30]. One genome assembly had Kleborate QC warnings due to ambiguous bases, genome size > 7.5 Mbp or < 4.5 Mbp, or N50< 10 Kbp and was removed from analysis. The COpy Number Variant quantifICation Tool (convict-v1.0) was used to identify antimicrobial resistance (AMR) from the ResFinder database (Accessed 2021-11-09) and estimate gene copy numbers as described previously (Shropshire, W convict GitHub: https://github.com/wshropshire/convict) [21]. CONVICT as part of its pipeline employs the KmerResistance-2.0 bioinformatic tool that uses short-read k-mer alignment (KMA) to identify homologous AMR genes from redundant databases [31, 32]. The NCBI AMRFinderPlus command-line tool (v3.11.14) was used to confirm AMR gene identification form CONVICT using the NCBI curated database (version 2023-04-17.1).

A short- and long-read assembly pipeline (Shropshire, W flye_hybrid_assembly_pipeline GitHub: https://github.com/wshropshire/flye_hybrid_assembly_pipeline) was used to close complete genomes of ONT sequenced data as described previously [21]. Incomplete assemblies were re-assembled using Unicycler-v0.5.0 and manually curated for errors using short- and long-read pileups and visualizing with the integrated genome browser (IGV-v2.14.1) [33]. The high-performance computing (HPC) cluster, Seadragon, that is hosted through MDACC was used to perform genomic analyses. Sequencing QC, 3GC-R gene determinants, and antimicrobial susceptibility testing for isolates sequenced for this project in addition to previous MDACC studies is available in Table S1.

### Core Gene and Genome Analyses of 3GC-R*Kp* Isolates

Genomes from index isolates (n=126) were annotated using Prokka-v1.14.5 and the annotation files in generalized feature format (GFF3) were used for the subsequent pan-genome analysis [34]. The gff3 files from Prokka were used to make a core gene alignment through Roary-v3.13 using MAFFT-v7.4 [35, 36]. The index 3GC-R*Kp* core gene length was 3720547 bases. The pairwise SNP distances from the core gene alignment were identified using snp-dists-v0.8.4 (https://github.com/tseemann/snp-dists). IQ-TREE-v2.0.6 was used to create a maximum likelihood phylogeny using the core gene alignment to determine the population structure [37]. Bootstrap analysis was performed using UFBoot approximation and SH-like approximate likelihood test with 1000 replicates though IQ-TREE-v2.0.6 [37, 38]. The tree visualization was performed using the R package ggtree-v3.9.1.

PopPUNK-v2.6.0 was used with index 3GC-R*Kp* draft assemblies (n=126) to identify potential cluster networks based on the core and accessory genome [39]. Following assignment, a core SNP phylogeny for each PopPUNK group was performed with a complete genome reference isolate using snippy-v4.6.0 (https://github.com/tseemann/snippy). The recombination regions were masked using Gubbins-v2.3.4 [36] and the filtered alignment file was used as the input file to obtain a core SNP maximum likelihood phylogeny using IQ-TREE-v2.0.6 [37]. The filtered polymorphic sites were used to generate pairwise SNP matrices to determine genetic relatedness within each specific PopPUNK group.

For CG307 population structure analysis, we performed a convenience sampling of publicly available CG307 isolates and oversampled for CG307 isolates harboring *bla*_SHV-205_ alleles. CG307 short-reads were downloaded from NCBI using the sratoolkit-v2.10.9 through the fasterq-dump function (https://github.com/ncbi/sra-tools). SRA accession numbers and metadata for CG307 isolates are available on Table S2. A core SNP phylogeny for the CG307 isolates was performed using Kp616 (GenBank Accession Number: GCA_003076555.1) as a reference isolate using snippy-v4.6.0 as described in the paragraphs above. The Kp616 isolate was selected as a reference as it was previously used by Wyres et al. for comparing CG307 population structure in addition to the genome being closed, completely resolved, and collected in 2009 [10]. The recombination regions were masked using Gubbins-v2.3.4 and a core SNP maximum likelihood phylogeny using IQ-TREE-v2.0.6 was performed as previously mentioned [37]. The per branch statistics output from Gubbins was analyzed to look for regions of high recombination.

A Bayesian analysis of population structure was performed using the core genome alignment through rhierBAPS-v1.0.1 to identify CG307 specific clusters[40]. Bayesian dating of the nodes of the CG307 phylogenetic tree was performed using BactDating-v1.1.0 (https://xavierdidelot.github.io/BactDating/). The recombination free core SNP phylogenetic tree output from Gubbins-v2.3.4 was used to perform root-to-tip regression analysis prior to molecular dating (Fig. S1). The output timed tree was further annotated and visualized using R package ggtree-v3.9.1.

### Accessory genome analysis

We used a subset of the accessory genome excluding low (<5%) and high frequency (>95%) genes for agglomerative hierarchical clustering of the gene presence/absence matrix from Roary to analyze the accessory genomes of CG307. Comparison of plasmid vectors with BLAST ring image generator (BRIG) was performed using the Proksee webserver [41]. Phage content in the genomes were characterized using the PHASTER webserver (https://phaster.ca/). Replicon types were identified using the “PlasmidFinder” database (Accessed 2023-07-01) with ABRicate-v1.0.0 (https://github.com/tseemann/abricate). Data visualization was generated using R-v4.0.4 packages or Geneious Prime software (2023.2.1).

### Statistical analysis

Time series data were assessed using the Mann-Kendall Trend test. Comparisons between rates of *K. pneumoniae* bacteremias by 6-month groups were performed using Student’s *t*-test. Chi-squared tests were used to compare group proportions between *K. pneumoniae* bacteremias. All statistics was computed on R-v4.0.4 or Prism 9 with a type one error rate (α=0.05) used across all hypothesis testing.

## RESULTS

### Identification of higher 3GC-R*Kp* bacteremia prevalence in latter half of annual periods

Out of 668 *K. pneumoniae* bacteremias that occurred from March 2016 to May 2022, 83% (554/668) were index infections (*i.e.*, the first occurrence of patient *K. pneumoniae* bacteremia episode). There were 114 (17%) index bacteremias with one or more recurrent infections (*i.e.*, bacteremia occurring ≥ 14 days following an index bacteremia episode). 3GC-R*Kp* infections accounted for 29% (194/668) of the *K. pneumoniae* bacteremias, of which 72% (139/194) were index infections. The proportion of index bacteremias that had recurrent episodes were significantly higher for 3GC-R*Kp* (39%, 55/139) compared to third generation cephalosporin susceptible *K. pneumoniae* (3GC-S*Kp*) infections (14%, 59/415, χ^2^ P-value < 0.001). When normalized by the number of admissions to account for fluctuations in patient volume, the rates of index 3GC-R*Kp* bacteremias were not significantly different over the course of the study (Mann Kendall trend test P-value > 0.05) (Fig. 1A). Similar to our recent study of 3GC-R *Escherichia coli* bacteremia [42], 3GC-R*Kp* bacteremia prevalence was significantly higher in the last six months of the year relative to the first six months (0.67 vs. 0.45 bacteremias per month per 1,000 patient admissions respectively; Student’s *t*-test P-value < 0.001) (Fig. 1B, 1C). No statistically significant difference in 3GC-S*Kp* bacteremias stratified by time of year was observed (1.6 vs. 1.3 bacteremias per month per 1,000 patient admissions in 2nd half of year vs. 1st half of year, respectively; Students *t*-test P-value = 0.07, Fig. 1C). These data indicate that for the past several years the rates of 3GC-R*Kp* bacteremia have not been significantly changing and that environmental factors may contribute to 3GC-R*Kp* infections.

**Fig. 1.**
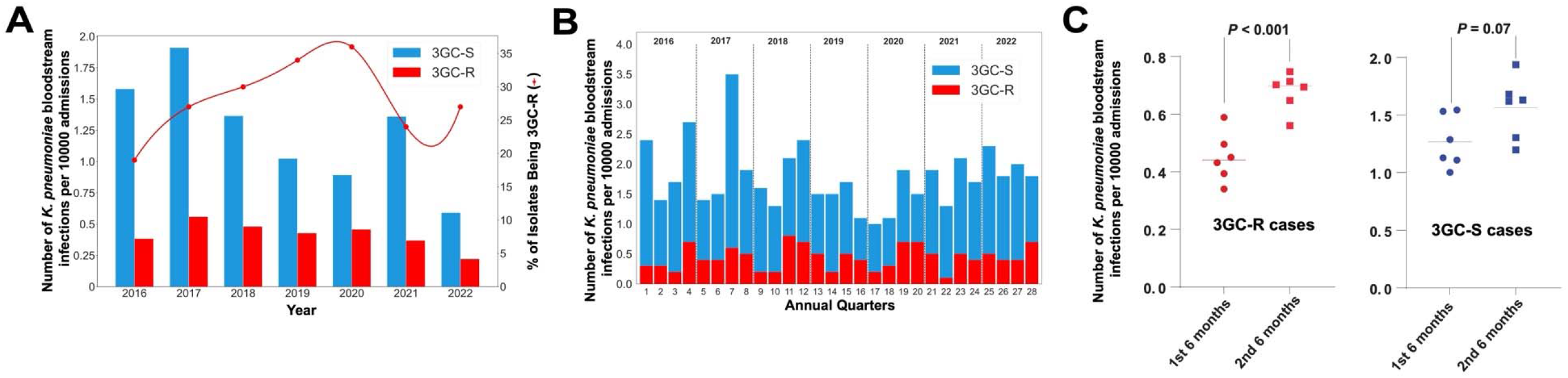
Epidemiology of *K. pneumoniae* bacteremias. (A) Index *K. pneumoniae* bacteremias by year. The x-axis shows the year, and the y-axis (left) shows the normalized number of bacteremias with blue bars representing the 3GC-S*Kp* and red bars representing the 3GC-R*Kp* infections, respectively. The y-axis (right) shows the proportion of 3GC-R*Kp* isolates amongst total *K. pneumoniae* bacteremias (red-line). (B) *K. pneumoniae* bacteremias stratified by quarters of the year. The x-axis represents quarters of the year, and the y-axis represents the normalized number of *K. pneumoniae* bacteremias with 3GC-R*Kp* (red) and 3GC-S*Kp* (blue) labelled respectively. (C) Comparison of 3GC-R*Kp* and 3GC-S*Kp* bacteremia in the first half and the second half of the year. The red dots on the left represent the average number 3GC-R*Kp* bacteremia for each month (*e.g.*, January, February, etc.), and the blue dots on the right represent the average number of 3GC-S*Kp* bacteremias by month. Student’s *t*-test P-values are labelled accordingly.

### CG307 strains caused more 3GC-R*Kp* bacteremias during study timeframe compared to other 3GC-R*Kp* genotypes

Out of the 194 3GC-R*Kp* causing bacteremia during the study period, there were 79% (161/194) 3GC-R*Kp* isolates available (Fig. 2A) that met our study’s inclusion criteria with 27 3GC-R*Kp* isolates included from a previous study [21]. One index 3GC-R*Kp* isolate had an assembly with genome size >7.5 Mbp and was excluded from the study. Further, we identified seven index 3GC-R*Kp* isolates (4%) that belonged to *K. pneumoniae* species complex taxa that were not *K. pneumoniae sensu stricto* and were thus excluded from analysis [4]. For the remainder of the study, all references to *K. pneumoniae* refer to *K. pneumoniae sensu stricto* strains. Thus, we had a total of 153 isolates with WGS data available for analysis with 126 index and 27 recurrent 3GC-R*Kp* isolates respectively (Fig. 2A). Among the 126 index isolates, there were a total of 52 unique STs detected highlighting the genomic diversity of our cohort. When grouping isolates by sharing at least five of seven loci from the Pasteur MLST scheme (*i.e.,* clonal group designations), a total of 19 unique CGs were identified (Fig. 2B). The most predominant clonal group identified from our index 3GC-R*Kp* isolates was CG307 (29%; 37/126) with all 37 isolates sharing identical ST307 schema. Interestingly, we only detected 6 index 3GC-R*Kp* isolates (5%) that belonged to the pandemic CG258 lineage with ST258 (n=2), ST11 (n=2), and ST395 (n=2) identified. Additional to CG258 and CG307 isolates, we only identified clonal groups with 5 or more index 3GC-R*Kp* isolates for CG29 (9/126; 7%), CG2947 (9/126; 7%), CG15 (8/126; 6%), and CG392 (6/126; 5%) with the remaining 40% (51/126) index 3GC-R*Kp* isolates belonging to rarely identified clonal groups (*i.e.,* <5 matching clonal group isolates detected). When stratified by full calendar years in our sampling frame (Fig. 2C), CG307 remained the most frequent CG detected per year except in 2021 when CG392 (n=4) was the most common CG detected (N.B., 2016 and 2022 were excluded due to incomplete annual sampling). We observed a statistically significant positive correlation (Pearson correlation coefficient *r*=0.91; P-value=0.03; Fig. 2D) between annual frequency of CG307 isolates collected and total 3GC-R*Kp* infections detected per year indicating that CG307 isolates may be the primary clonal group contributing to 3GC-R*Kp* bacteremia prevalence.

**Fig. 2.**
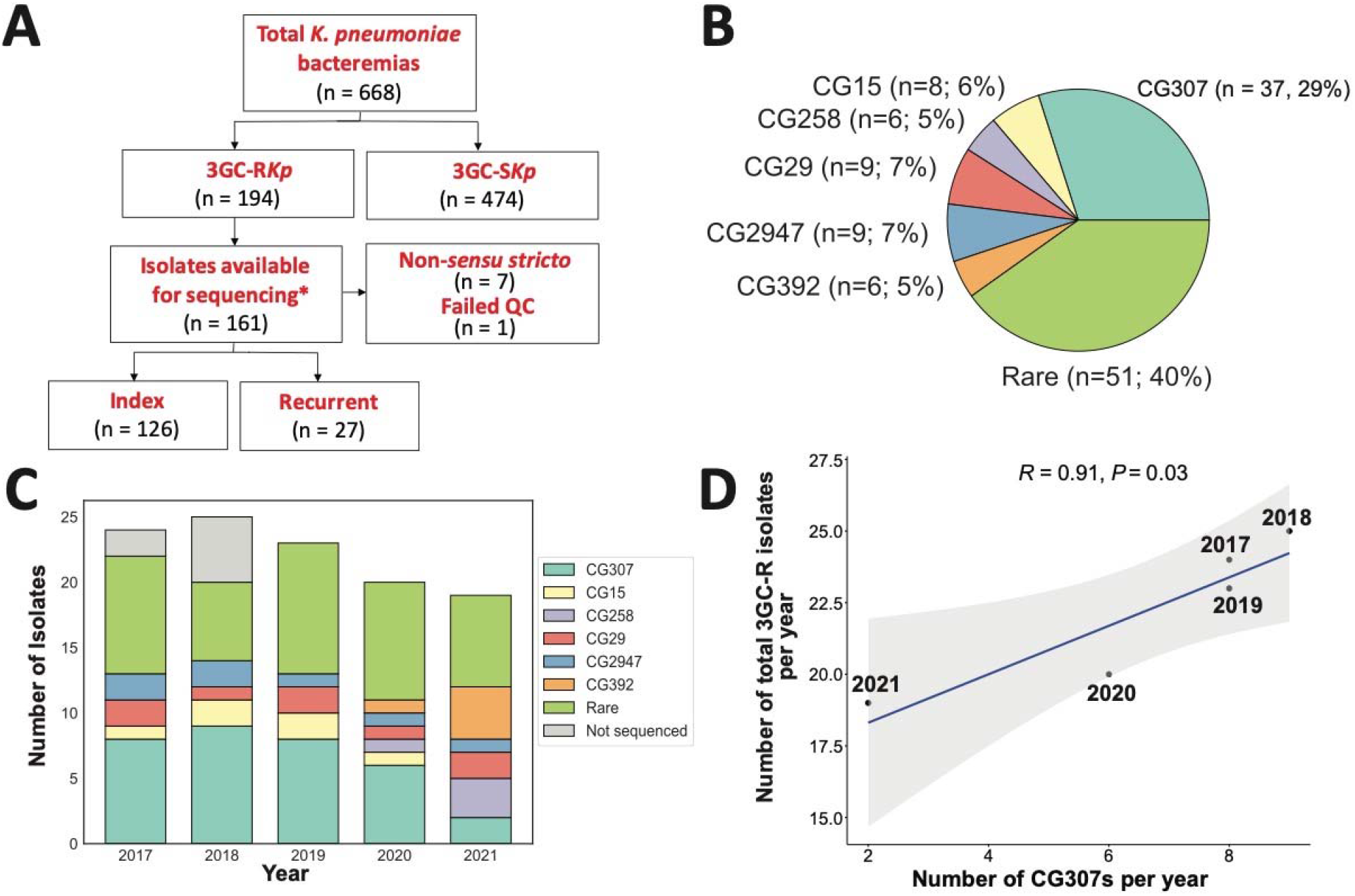
Whole genome sequencing workflow, clonal group distributions, and trends of 3GC-R*Kp* isolates. (A) Workflow of WGS inclusion/exclusion criteria for *K. pneumoniae* bacteremia isolates. *27 isolates from a previous study (BioProject Accession #: PRJNA836696) are included in our analyses. (B) Pie chart showing the clonal group distribution of index 3GC-R*Kp* isolates. (C) Clonal groups of index 3GC-R*Kp* isolates stratified by year. Legend indicates clonal groups by color with gray representing index 3GC-R*Kp* isolates that were not sequenced. (D) Positive correlation between sequenced CG307 isolates per year (x-axis) and the total number of index 3GC-R*Kp* isolates from 2017-2021 (y-axis). Pearson’s correlation coefficient (*r*) with associated correlation test P-value reported in figure.

### High genomic diversity within index 3GC-R*Kp* core gene population structure

To gain further insights into 3GC-R*Kp* population structure, we performed a maximum-likelihood phylogeny inferred by a core gene alignment of index 3GC-R*Kp* isolates (n=126) and identified diverse, deep-branching lineages often observed in MDR *K. pneumoniae* [4]. Using a core gene threshold definition of gene presence in ≥99% of cohort, 16% (3873/24209) of the pangenome was ‘core’ gene content reflecting substantial pangenome diversity. The median pairwise nucleotide divergence was 0.58% (median pairwise SNP distance [MPSD] = 21402) comparable to previous estimates [3, 20]. Although most CGs evidenced significant genetic diversity, there were clear clusters of closely related CG307 (MPSD = 84) and CG29 (MPSD = 15) isolates (Fig. 3). Both the CG307 (KL102) and CG29 (KL19) isolates had highly conserved capsule locus (KL) genes compared to the other CGs (Fig. 3). Similar to the KL locus, the O-locus was highly conserved in CG307s (O2v2), CG29 (O1v2), and CG15 (O1v1) when compared to the other CGs. When analyzing the primary determinants of hypervirulent *K. pneumoniae* (hvKP) [29, 43], the siderophore yersiniabactin locus (*ybt*) was present in 31% of our 3GC-R*Kp* population, consistent with the findings of Lam et al. 2018 [43]. Less commonly detected hvKP determinants were the genotoxin colibactin (*clb*, n=4), siderophore aerobactin (*iuc*, n=4), siderophore salmochelin (*iro*, n=1), and hypermucoidy *rmpADC* operon (n=1). One particularly interesting isolate belonged to ST412 (*i.e.,* MB3190), which is a member of hypervirulent-associated CG23, that harbored the *rmpADC* operon in addition to *iro1* and *iuc1*, often carried on the FIB_k_ virulence plasmid KpVP-1 [43].

**Fig. 3.**
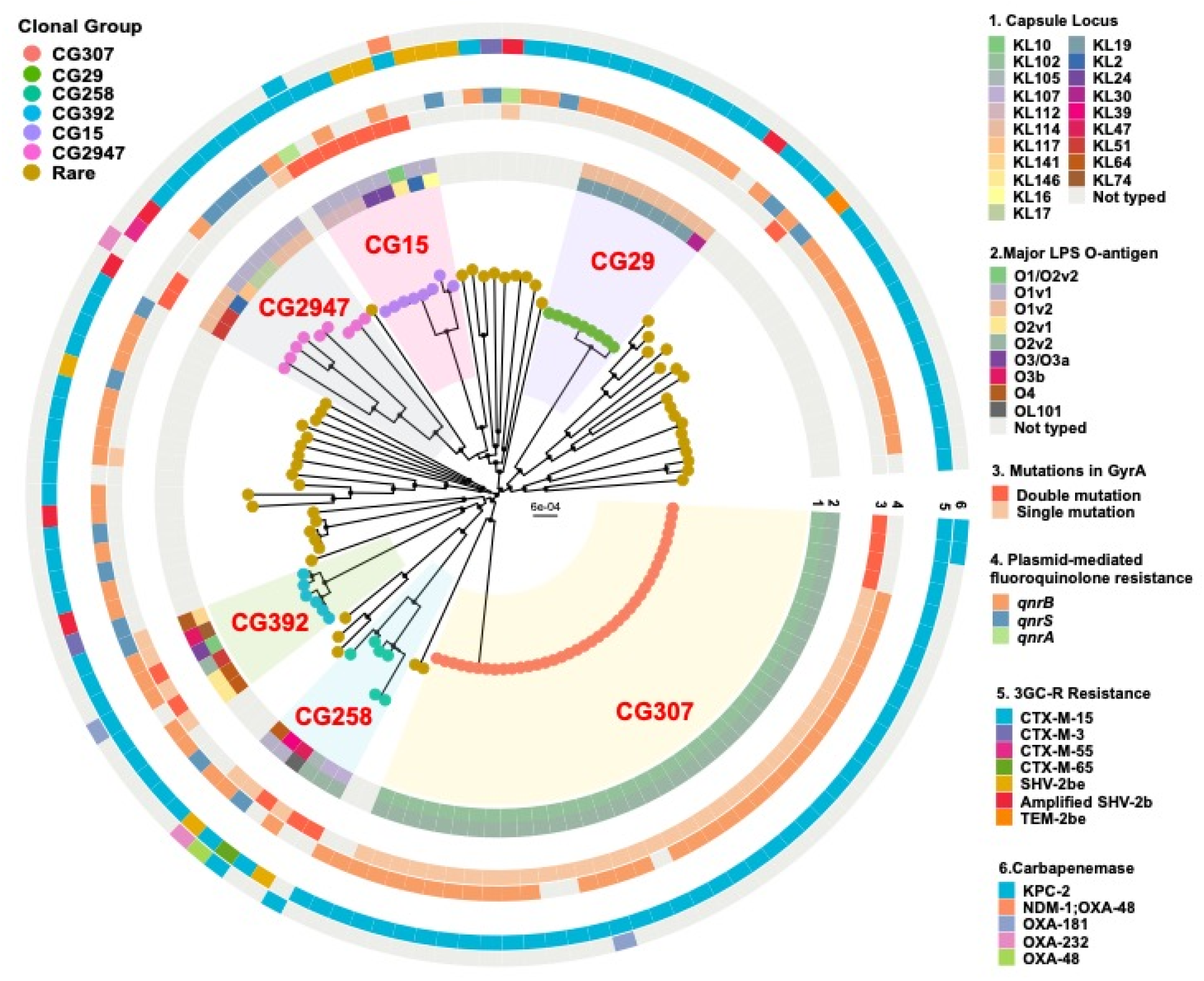
Core gene maximum-likelihood inferred phylogenetic analysis of index 3GC-R*Kp* strains. Phylogenetic tree is midpoint rooted with circular internal nodes representing 95% UFBoot confidence values. Branch tips are labelled by clonal group with each CG highlighted within tree structure. Rings are labelled as follows: 1) Capsule type; 2) O-antigen; 3) GyrA polymorphisms; 4) Plasmid-mediated fluoroquinolone resistance genes; 5) 3GC-R resistance genes; and 6) Carbapenemase encoding genes.

When focusing on the genetic determinants associated with 3GC-R*Kp*, 94% (119/126) harbored AMR genes encoding enzymes that are associated with a 3GC-R*Kp* phenotype. The overwhelming majority of 3GC-R*Kp* isolates (n=109) harbored the ESBL encoding gene, *bla*_CTX-M_ with 95% (104/109) harboring the *bla*_CTX-M-15_ variant. We detected *bla*_SHV-2be_ variants in eight strains, only two of which were identified using the NCBI AMRFinderPlus database with our genomic assemblies; the remainder required the use of a short-read, k-mer mapping to database approach with KmerResistance to identify homologous *bla*_SHV-2b_ and *bla*_SHV-2be_ variants in the same strains. Additionally, CONVICT identified five strains with amplification of *bla*_SHV-2b_ variants (*i.e.*, gene copy number of ≥ 2.0×), with similar amplifications of penicillinase encoding genes associated with a ‘false ESBL phenotype’ [44]. A carbapenemase encoding gene was observed in 11 strains (9%) (*bla*_KPC-2_, n =5; *bla*_OXA-48_, n=2; *bla*_OXA-181_, n=2; *bla*_OXA-232_, n=2) (Fig. 3). In total, a putative genetic mechanism for the 3GC-R phenotype was present in 125/126 index 3GC-R*Kp* isolates (99%).

Given the extensive use of fluoroquinolones (FQs) as prophylaxis for neutropenic patients at our institution in addition to associated FQ resistance (FQR) and ESBL producing Gram-negative organisms [45], we next analyzed fluoroquinolone susceptibility and FQR mechanisms. Ciprofloxacin non-susceptibility was observed in 73% (92/126) of index 3GC-R*Kp* isolates. Quinolone resistance determining region (QRDR) mutations in *gyrA* and *parC* were observed in 50% (64/126) and 41% (52/126) of the strains, respectively, whereas the most commonly observed FQR mechanism in our cohort was the plasmid mediated FQR gene *qnr,* primarily *qnrB1*, observed in 79% (101/126) of the strains. Single and double QRDR mutation for *gyrA* were almost exclusively observed in the major CGs such as CG307, CG258, CG392, and CG15, whereas rare CGs typically lacked *gyrA* QRDR mutations but did contain *qnr* genes (Fig. 3). The double mutation pattern *gyrA-83+87* was present in 14 strains including four CG307s (Fig. 3).

### 3GC-RKp transmission and recurrence dynamics driven primarily by CG307 and CG29 clades

Given the known capacity of 3GC-R*Kp* to be transmitted in healthcare settings [2, 46], we used PopPUNK to identify nested populations of 3GC-R*Kp* and subsequently performed PopPUNK cluster specific, core genome alignment inferred phylogenies masked for recombination to potentially identify transmission networks. We were able to identify 56 PopPUNK groups of which 14 included two or more isolates (*i.e.*, 42 isolates were uniquely divergent from the full cohort, consistent with our clonal group population structure). From these 14 PopPUNK groups, we identified 28 3GC-R*Kp* isolates from unique patients that differed by <25 pairwise SNPs (*i.e.,* a SNP threshold that has been previously used to define potential *K. pneumoniae* transmission [47]) with a minumum of one other unique patient 3GC-R*Kp* (Table S3). Potential strain transmission cluster sizes varied from 2-8 isolates with a median pairwise SNP distance of 8 and range of 0-24 SNPs. Putative transmission clusters with genetically related pairwise isolates involved four distinct STs, namely ST307 (five clusters, 16 total isolates), ST29 (one cluster, eight isolates), ST152 (one cluster, 2 isolates), and ST280 (one cluster, 2 isolates). Interestingly, 97% (27/28) of index isolates from putative transmission clusters caused infections in patients with hematologic maligancy compared to 66% (65/98) non-clustered strains (χ^2^ P-value < 0.001). The two largest transmission clusters (ST307 [pp8-1], n = 7; ST29, n = 8 [pp65-1]) both occurred over prolonged periods of time, 28 months for ST307 pp8-1 cluster and nearly 60 months for the ST29 pp65-1 cluster in patients with multiple admissions and extensive contact with the healthcare system (Fig. 4A).

**Fig. 4.**
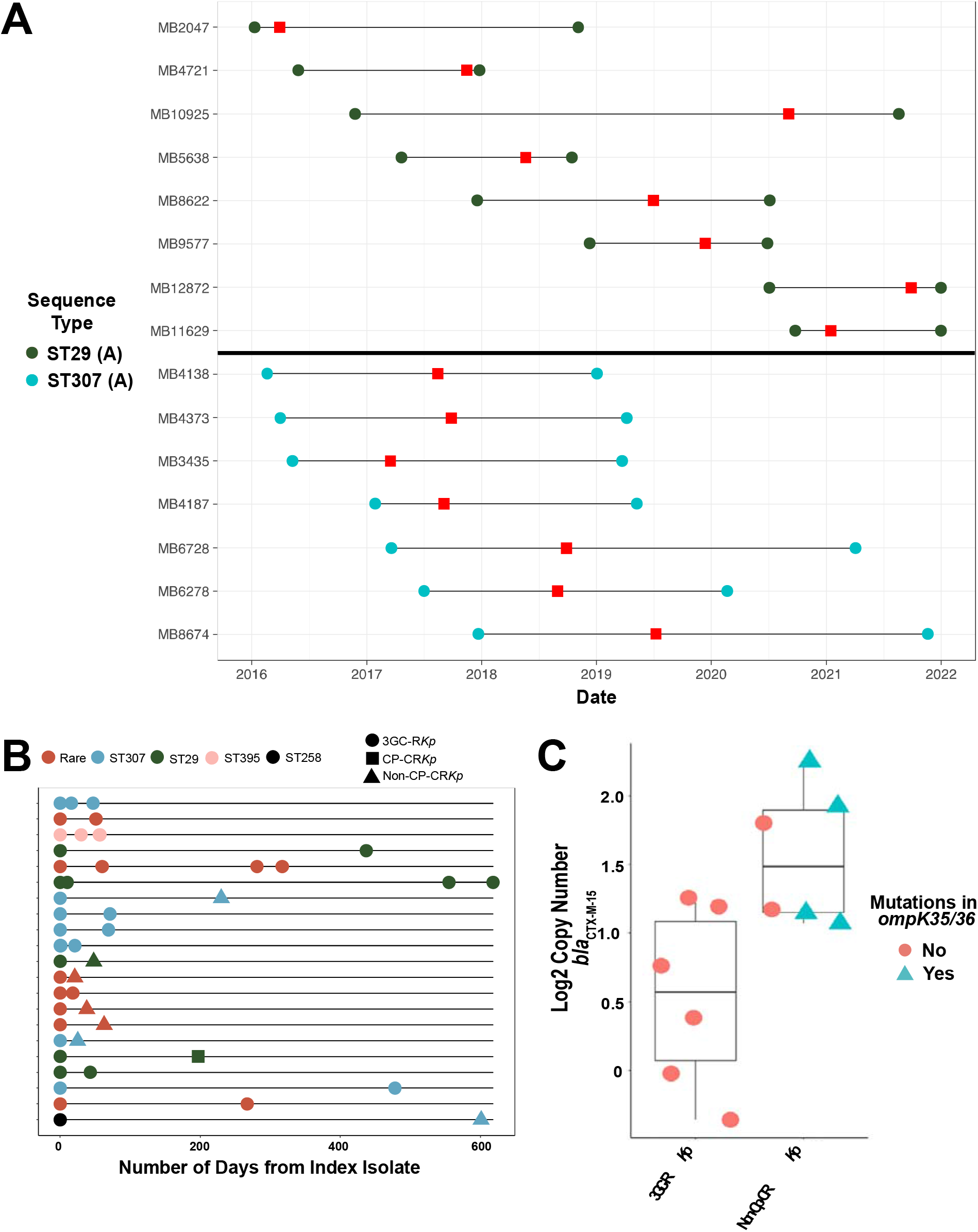
3GC-R*Kp* transmission and recurrence dynamics. (A) Genetically related (*i.e.*, <25 pairwise SNPs) isolates collected over time from unique patients for CG29 (top; dark green; transmission cluster pp8-1) and CG307 (bottom; light blue; transmission cluster pp65-1) clades. Circles represent start and end of healthcare interactions at our institution for each respective patient with horizontal black line representing the duration of healthcare services. Red rectangles indicate collection date for each respective unique isolate. Note that patient-sample pair for MB12872 and MB11629 had end dates that went past 2021/12/31. (B) Index 3GC-R*Kp* isolates with paired recurrent (*i.e.*, *K. pneumoniae* bacteremia isolate collected >14 days from antecedent index isolate). Duration of days from index to recurrent isolate is shown on the x-axis. Colors denote sequence type and shape indicates phenotypes as shown in legend. 3GC-R*Kp* = third-generation cephalosporin resistant *K. pneumoniae*; CP-CR*Kp* = carbapenemase producing carbapenem-resistant *K. pneumoniae*; Non-CP-CR*Kp* = non-carbapenemase producing carbapenem-resistant *K. pneumoniae* (C) *bla*_CTX-M_ copy numbers and *ompK35/ompK36* mutation status in paired 3GC-R*Kp* (left) and recurrent Non-CP-CR*Kp* (right) isolates. Y-axis shows log2 transformed *bla*_CTX-M-15_ copy number estimates. Shapes and colors show *ompK35/ompK36* status as delineated in the legend.

During the study timeframe, 12% (21/126) of index *b*acteremia patients had a recurrent 3GC-R*Kp* bacteremia occurring an average of 94 days following the initial 3GC-R*Kp* bacteremia episode (Fig. 4B). The same ST as the index isolate caused 91% of the recurrent infections. Consistent with carbapenems being the primary treatment for 3GC-R*Kp*, 38% (8/21) of isolates causing recurrent infections developed carbapenem resistance (Fig. 4B). The majority of strains (n=7) developed carbapenem resistance in the absence of a carbapenemase gene (*i.e.*, were non-carbapenemase producing carbapenem resistant *K. pneumoniae* or non-CP-CR*Kp*), whereas a single ST29 strain acquired *bla*_KPC-2_ (Fig. 4BB). Through a combination of ONT long-read sequencing in addition to CONVICT gene copy number estimates, we found that six out of the seven non-CP-CRE recurrent isolates had evidence of *bla*_CTX-M-15_ gene amplification (Fig. 4C). Additionally, mutations in *ompK*35 (n=1) and *ompK*36 (n=4) were observed in 5/7 (71%) of the non-CP-CR*Kp* isolates (Fig. 4C). These data indicate that recurrence of 3GC-R*Kp* bacteremia primarily occurs due to re-infection by the same strain which has often developed carbapenem resistance through non-carbapenemase mechanisms.

### Identification of a novel lineage of CG307 causing bacteremia at our institution

Given the stable detection of CG307 within the Houston region [7, 9, 20], including previously at our own institution [21], we further analyzed CG307 isolates from our cohort (n=37) along with a worldwide distribution of geographically diverse CG307 isolates (n=187) publicly available on NCBI. A Bayesian dated, recombination masked, phylogeny of 224 CG307s inferred from a core genome SNP alignment is presented on Fig. 5. Initial root-to-tip analysis of our CG307 cohort indicated a strong temporal signal (Fig. S1). Consistent with previous data, the CG307 splits into two primary clades consistent with the previously described “Texas-specific” (n =74) and “global” clades (n = 150). Within these respective CG307 clades, there were two nested sub-populations within each clade with clusters 3 and 4 belonging to the Texas-specific lineage and clusters 1 and 2 as part of the Global lineage [10, 20]. The Texas-specific clade harbored double GyrA-83I-87N mutations and 2× copies of *bla*_CTX-M-15_ that have previously been characterized (Fig. 5) [10]. Interestingly, only four CG307 isolates in our cohort clustered with the Texas-specific clade. In contrast, the majority (n=33) of our CG307 isolates belonged to the global clade with an approximate even split between nested populations (18 and 15 isolates in clusters 1 and 2 respectively).

**Fig. 5.**
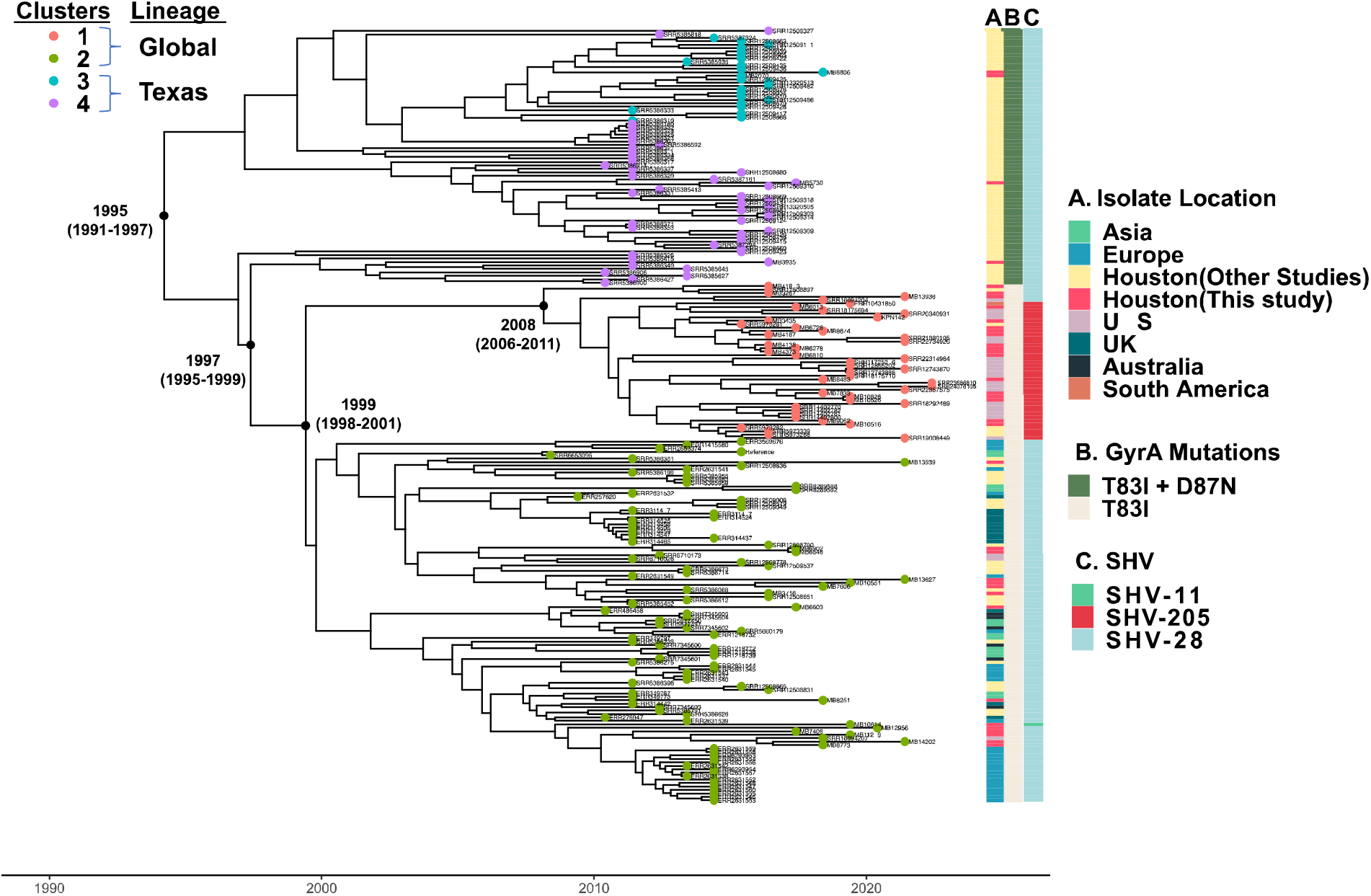
Identification of unique CG307 sub-clade (*i.e.,* cluster 1) causing bacteremia at our institution. Presented is a Bayesian dated, core SNP recombination masked, phylogeny of 224 CG307 strains using isolate Kp616 (BioSample Accession Number: SAMN08391417) as reference. Tip colors indicate hierBAPS predicted taxa with clusters 1 and 2 belonging to the previously identified global clade and clusters 3 and 4 being part of the Texas-specific clade. Years indicate predicted dates of divergence from internal nodes of interest as estimated using BactDating. Dates in parentheticals represent the 95% confidence interval. Metadata presented on right columns include A) geographic location of isolate collection; B) presence of *gyrA* mutations; and C) chromosomal *bla*_SHV_ isoforms.

Our Bayesian dating analysis of our phylogeny estimates CG307 emerged in 1995 (95% confidence interval [95%CI]: 1991 – 1997), consistent with a previous estimation of CG307 emergence in 1994 [10]. The date of divergence between the Texas-specific and global CG307 clades is estimated to have occurred in 1997 (95%CI: 1995 – 2000) shortly after CG307 had emerged, suggesting that CG307 may have originated from the Houston region as previously hypothesized [10]. When focusing on the global clade, cluster 1 strains were estimated to emerge from cluster 2 strains approximately 1999 (95%CI: 1998-2001) with cluster 1 isolates beginning to clonally expand near 2008 (95%CI: 2006-2011) (Fig. 5). A conserved genomic feature of cluster 1 CG307 strains was the stable vertical transmission of *bla*_SHV-205_, a single amino acid variant of the chromosomal *bla*_SHV-1_ gene (Fig. 5). Consistent with our estimation of cluster 1 strains emerging near 2008, all publicly available cluster 1 strains with metadata were isolated from 2016 onwards with 15/25 isolated since 2020. All but one strain (Paraguay 2020) came from the United States including such geographically diverse locales as Utah, Chicago, and New York and even included two strains isolated from dogs in Texas. Given that MDA has a broad geographic patient distribution, we analyzed the home location for each patient with a CG307 infection at our institution and found that regardless of the cluster of infecting isolates, most patients (31/37, 82%) were from Texas and the majority were from the Houston area (54%) (Fig. S1A). There were three patients from the United States who resided outside Texas all of whom were infected by strains of the global lineage as were the three patients from outside the United States (Fig. S1B). These data indicate that the global lineage of CG307 caused the majority of CG307 infections even among our patients from Houston and the rest of Texas.

### Differing accessory genome content across each of the CG307 clades

To gain further insight into the various CG307 clusters, we used our Illumina data in conjunction with ONT long-read sequencing to dissect accessory genome content and determine genomic context of 3GC-R*Kp* determinants. We subset all accessory genome content shared in greater than 5% but less than 95% of the CG307 population to look for gene presence/absence signals present across our four clades and observed a demarcation across the four clusters (Fig. 6A). We noted that there were global and Texas-specific clade differences in replicon type detection, which has been described in previous literature (Fig. 6B) [20]. The global clade had an enrichment of IncFII (84%) and IncFIB_K_ (97%) whereas there were very few cluster 3 isolates with either replicon detected (IncFII [36%]; IncFIB_K_ [20%]) (Fig. 6B). These multireplicon F-type plasmid differences correlates with *bla*_CTX-M-15_ carriage differences between global and Texas-specific clades as previously described [10, 20]. As shown in Fig. S3A, both MB5730 (Cluster 4) and MB8806 (Cluster 3) from the Texas-specific clade harbored 2 copies of *bla*CTX-M-15 inserted in the chromosome. Conversely, isolates MB7606 (Cluster 2) and MB8674 (Cluster 1) from the global clade harbored *bla*CTX-M-15 in association with IS*26* pseudo-compound transposons made of two or more IS*26* units with directly flanking IS*26* transposases on IncFIB_K_ plasmids (Fig. S3B).

**Fig. 6.**
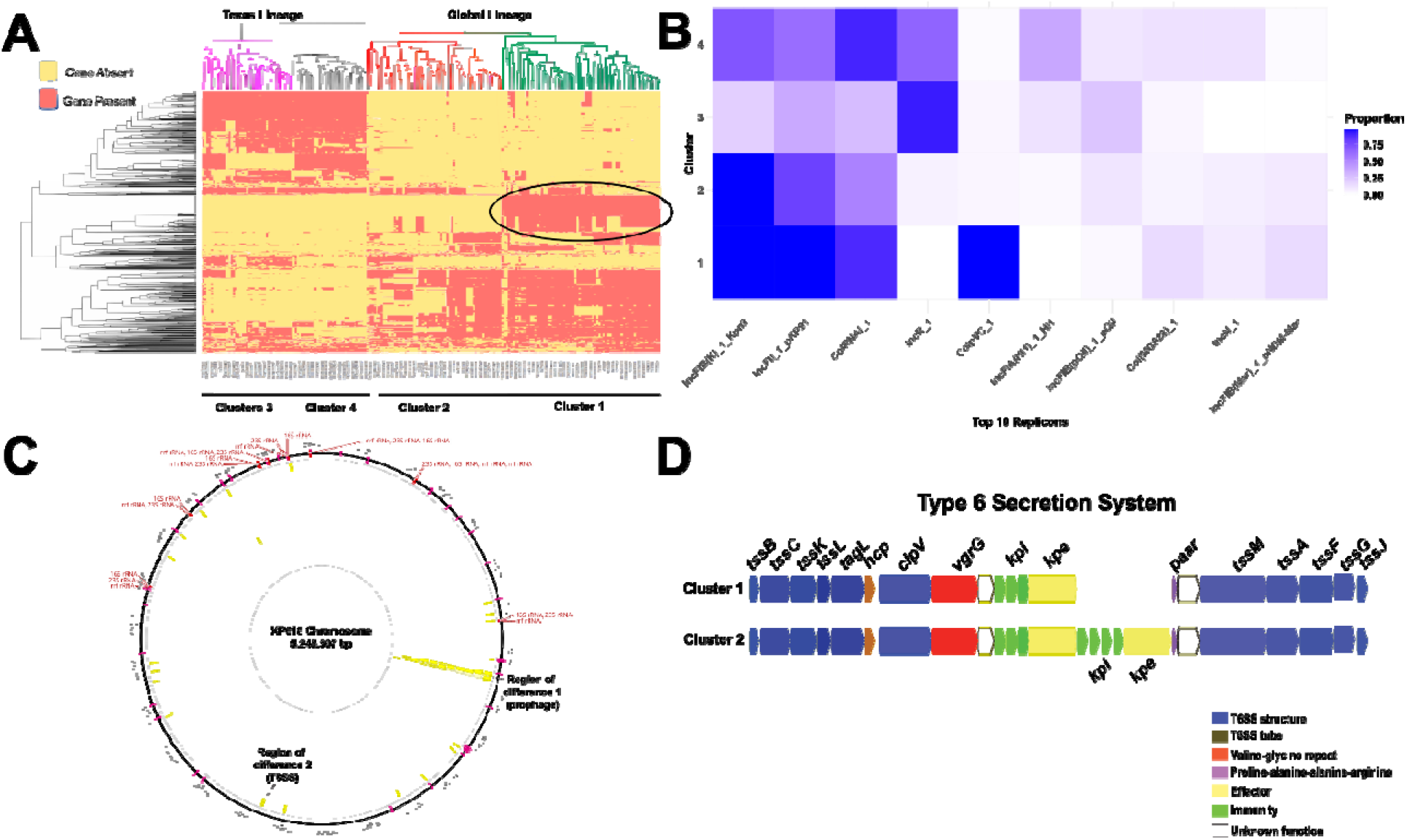
Assessment of accessory genome differences between CG307 strains. (A) Gene presence/absence heatmap representing the agglomerative hierarchical clustering of accessory genomes of CG307 strains. Circled region indicates genetic content unique to cluster 1 isolates. (B) Proportion heatmap of the most common replicon types identified using PlasmidFinder stratified by CG307 cluster. (C) Recombination hotspots (yellow blocks) of CG307 isolates using Kp616 as a reference and Gubbins predicted recombination sites output. Regions labelled on chromosome indicate genomic context we identified that differed between cluster 1 and 2. (D) Region of difference 2 between CG307 cluster 1 and cluster 2 strains involving a type 6 secretion system (T6SS). Labeling system for T6SS proteins is taken from a previous study [48] with description of color of putative gene function shown in the legend. Note that cluster 1 strains lack of a set of immunity/effector proteins present in cluster 2 strains.

We focused on accessory genome differences between the global cluster 1 and cluster 2 isolates to determine what was driving the potential emergence of cluster 1 in our region. We found that cluster 1 strains had a very high recombination to mutation (r/m) ratio of 16 suggesting that recombination events were driving diversity in this clade. We found two recombination blocks in cluster 1 isolates that appeared to be regions that differed with cluster 2 isolates (Fig. 6C). First, we found that, relative to cluster 2 strains, cluster 1 strains uniquely contain a 40 Kbp prophage-like region which contains numerous putative regulatory proteins as well as proteins of unknown function (Fig. 6C). Second, although both cluster 1 and cluster 2 strains contain a type 6 secretion system (T6SS), cluster 1 strains lack a set of T6SS effector/immunity proteins which are present in cluster 2 strains (Fig. 6D). Taken together, we conclude that the cluster 1 strains identified in our study are a distinct genetic branch of the global CG307 clade that have unique accessory gene content as well as are causing disease throughout the United States.

## DISCUSSION

*K. pneumoniae* is a major contributor to healthcare associated infections and can acquire resistance to a diverse array of antimicrobials thereby rendering treatment problematic [1, 50, 51]. Given the limited molecular epidemiology of 3^rd^ generation cephalosporin resistant *K. pneumoniae* (3GC-R*Kp*) in the United States, we characterized 126 index and 27 recurrent 3GC-R*Kp* bacteremia isolates collected from 2016 to 2022 at our institution (Fig. 2A). Consistent with other worldwide MDR *K. pneumoniae* surveillance studies, we observed long-branching, genetically diverse 3GC-R*Kp* isolates [22, 23, 47, 52-55]; however, there were clusters of “global problem clones” including CG307, CG29, and CG15 isolates with evidence of limited transmission [4]. Moreover, through analysis of our patients and publicly available genomes, we have identified a previously uncharacterized *K. pneumoniae* CG307 sub-clade (*i.e.,* cluster 1) that caused 14% of our index 3GC-R*Kp* infections and has been isolated recently from multiple locations across the United States [26, 55].

A major impetus for our study were reports of a recent expansion of 3GC-R*Kp* in the United States [12, 56]; however, prospective data from active 3GC-R*Kp* surveillance is limited, and thus increases in incident 3GC-R*Kp* infections remain unclear [57]. During our study timeframe, we observed fairly stable absolute frequencies as well as admission adjusted prevalence of 3GC-R*Kp* bacteremias (Fig 1). It has been suggested that increases in 3GC-R *Enterobacterales* infections in the United States have been largely driven by community-onset cases [50]. The genetic diversity of 3GC-R*Kp* isolates (Fig. 3) in conjunction with seasonal prevalence differences of 3GC-R*Kp* infections (Figs. 1B, 1C) suggest a possible community-wide dissemination in our region. These data are similar to those recently described for a hospital network in Australia in which comprehensive WGS of *K. pneumoniae* found that the majority of infections were caused by widespread circulation in the community rather than hospital transmission [47]. Consistent with our observation 3GC-R*Kp* infections tended to occur in the warmer second half of the calendar year compared to the first half of the year, a multi-site surveillance study across several continents found that environmental factors associated with warmer months may increase incidence rates of *K. pneumoniae* infections [58]. Nevertheless, similar to a recent analysis on 3GC-R *E. coli* prevalence at our institution [42], we observed a statistically significant association only for 3GC-R*Kp* but not 3GC-S*Kp* infections. Thus, our results taken together with previous studies suggest that limiting 3GC-R*Kp* acquisition outside of the hospital is likely to be critical to mitigation efforts.

Although we had a long-branching, genetically diverse cohort of 3GC-R*Kp* isolates (Figs. 2, 3), we also observed numerous instances of potential hospital-based 3GC-R*Kp* transmission (Fig. 4A). It is worth noting that given we only sampled 3GC-R*Kp* isolates collected from bacteremia infections, we are likely underestimating the true scope of transmission that could be occurring at our institution. Interestingly, transmission clusters almost exclusively were due to the “problem clones” CG307 and CG29 (Fig. 4A) and involved patients with hematologic malignancy, likely because of the propensity of such individuals to develop bacteremias following initial colonization [59]. Outbreaks of these two “global problem clones” have been described [60] although whether such transmission is happening due to direct human contact or being acquired from the hospital environment is not currently known. Interestingly, for both CG307 and CG29 strains, we observed highly genetically related bacteria causing infections over a several years’ time-frame (Fig. 4A), suggesting potential environmental sources as has been observed for *Pseudomonas* spp. [61] and vancomycin resistant *Enterococcus faecium* [62]. The lack of clear epidemiologic links between infected patients means that standard infection control approaches would be unlikely to detect such transmission events emphasizing the potential complementary role that WGS could play in infection control efforts if real-time data were available [63].

Recurrence is a major concern following treatment of 3GC-R*Kp* infections [64, 65], and indeed we observed an approximate 20% recurrence rate when just considering bacteremias. The recurrent strain was nearly always the same ST and highly genetically related to the index isolate suggesting re-infection rather than acquisition of a new 3GC-R*Kp* strain. Interestingly, the recurrent organism could either retain a 3GC-R, carbapenem-susceptible phenotype or transition to carbapenem resistance, usually without acquisition of a carbapenemase (Fig. 4B). Although data are limited, a few studies have found that intravenous carbapenem administration does not reliably eradicate ESBL positive *Enterobacterales* from the intestinal tract, presumably due to low carbapenem stool concentrations [66, 67]. Alternatively, it has been recently demonstrated that *Enterobacter* spp. can enter into a cell-well deficient spheroplast capable of surviving carbapenem exposure and returning to normal growth once carbapenem pressure is removed, and thus carbapenem “tolerance” could have accounted for strain persistence [68]. 3GC-R*Kp* isolates can also develop outright resistance to carbapenems in the absence of carbapenemase by limiting drug influx through porin mutations along with hyperproduction of *bla*_CTX-M_ encoding enzymes, which we were able to demonstrate in multiple serial isolates using a long-read sequencing approach [21]. Remarkably, we collected many recurrent isolates months or years from the index isolate collection date (Fig. 4B) with minimal pairwise SNP differences (*i.e.,* < 10 SNPs), suggesting the long duration of colonization achievable by 3GC-R*Kp* [69]. We envision that a strategy of targeted *K. pneumoniae* gastrointestinal tract decolonization, such as using CRISPR-based technologies, could be part of the future treatment of 3GC-R*Kp* infections [70].

We were intrigued by the paucity of Texas-specific CG307 isolates in contrast to the Global CG307 clade collected in our cohort in addition to the relatively low number of CG258 isolates detected (Fig. 2). A study from a Houston-wide hospital system analyzing ESBL *K. pneumoniae* strains collected from 2011-2015 found similar numbers of CG307 and CG258 strains, an unexpected result given the predominant prevalence of CG258 in most United States investigations [5, 9, 19]. A comparative genomics analysis of CG307 isolates found that the Texas-specific clade was genetically distinct from other CG307 strains circulating worldwide, with multiple chromosomal insertions of *bla*_CTX-M-15_ and a fixed double GyrA mutation as a predominant genomic feature of the Texas-specific CG307 clade [10]. Another Houston based study found divergent accessory genomes between CG258 and CG307 with greater plasmid content in the latter, suggesting that CG307 isolates had potential greater capacity to adapt to selective pressures [20]. Nevertheless, this study found very few of the global CG307 clade circulating in their particular hospital system [20]. The reason for our finding that only a small fraction of CG307 isolates belong to the Texas lineage is not clear but might be due to temporal variation given our most recent sampling timeframe. Support for this hypothesis comes from the relatively recent isolation of CG307 strains from across multiple United States locales including Central Texas [26] and Mississippi that cluster together with the majority of our Global CG307 isolates. These isolates have a distinct chromosomal SHV, lack the multiple copies of chromosomal *bla*_CTX-M-15_ hypothesized to be important to the success of the Texas-specific lineage, and contain a unique accessory genome relative to other CG307 strains (Figs. 5, 6). Our findings mirror similar, recent clonal expansions of CG307 strains from the global lineage recently observed in Wales and Norway, suggesting that the United States may be in the early stages of dissemination of a CG307 Global sub-clade [22, 23].

We identified two major events distinguishing the Global CG307 cluster 1 and cluster 2 isolates. The cluster 1 strains contained a prophage that did not contain any clear virulence factors, although numerous open reading frames encoded proteins of unknown function and acquisition of exogenous DNA has been shown to influence the transcriptome of distant genes possibly through the presence of transcriptional regulators in the acquired DNA [71, 72]. Additionally, relative to cluster 1, cluster 2 strains had a distinct set of effector/immunity proteins from a type 6 secretion system (Fig. 6D). T6SS function as injection machinery that can directly puncture both eukaryotic and prokaryotic membranes and have been identified as contributing to *K. pneumoniae* pulmonary infection, gastrointestinal colonization, bacterial competition, and liver abscess formation [73-76]. Future work will be necessary to further functionally characterize potential differences in virulence and resistance factors contributing to each CG307 clades’ respective success.

Although our study had many strengths, there are some limitations. First, our isolates were recovered from a single center and thus the generalizability of our findings are not known; however, our cancer center draws from a large geographic region, and Houston has been a notable center of 3GC-R*Kp* infections [9, 20]. Second, we only analyzed bloodstream infections, which limits our results’ generalizability when it comes to other 3GC-R*Kp* infectious sources; nevertheless, given the invasive nature of bacteremia, this reduces likelihood of identifying false positive infections. Finally, the genetic diversity of 3GC-R*Kp* isolates meant that we could only analyze in-depth a few clonal groups even with a sufficient large sample size of over 150 3GC-R*Kp* samples sequenced.

In summary, we present a contemporary, comparative genomics-based analysis of 3GC-R*Kp* causing invasive disease in immunocompromised patients. We found that a previously unrecognized CG307 sub-clade is causing significant disease both in Houston and in diverse locations across the United States suggesting ongoing spread of an MDR *K. pneumoniae* strain with unique accessory genome content. Our findings emphasize the continued need for 3GC-R *Enterobacterales* surveillance in addition to the efforts made to track carbapenem resistant strains.

**Fig. S1.**
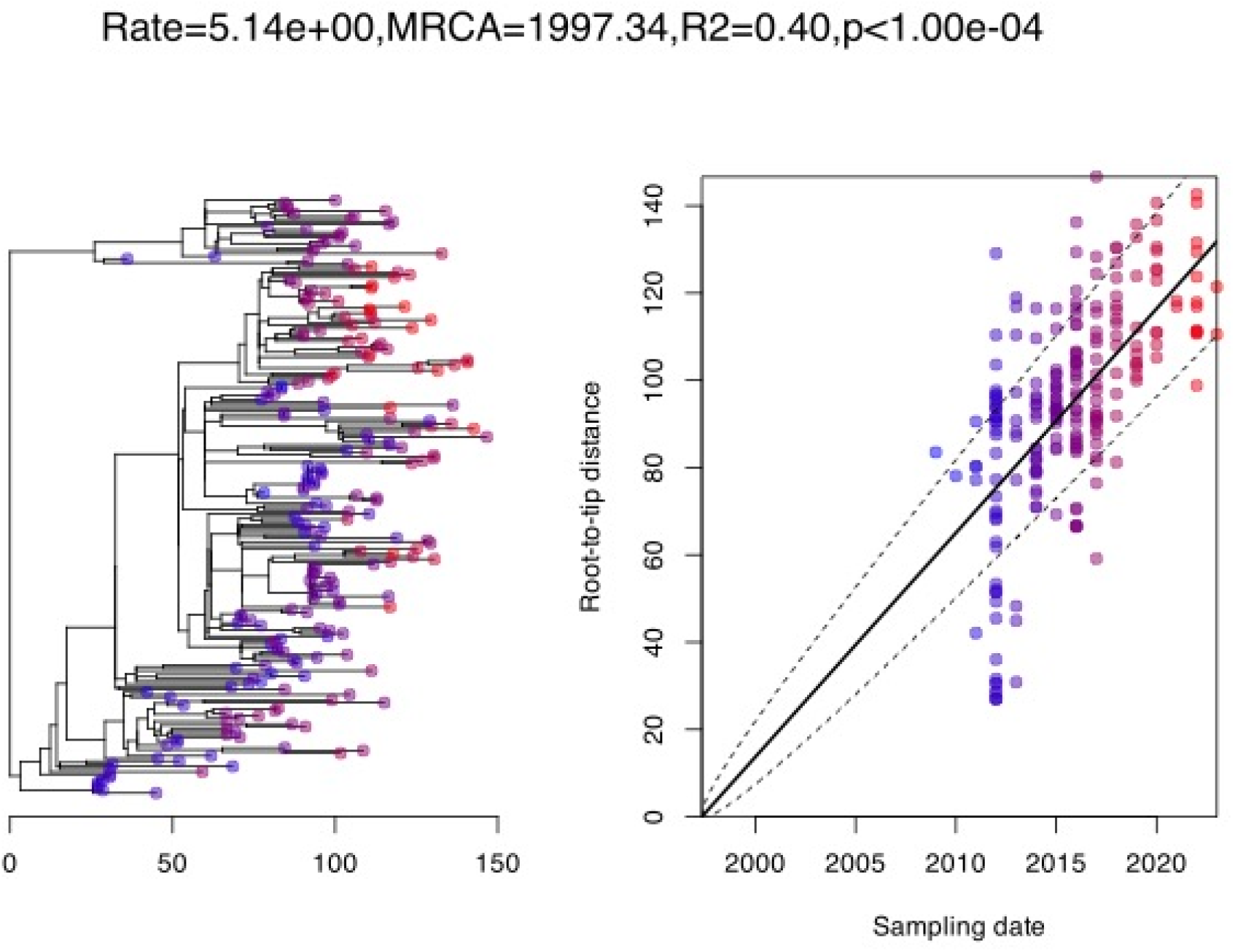
Initial root-to-tip correlation analysis of 224 CG307 isolates. Initial test of Gubbins phylogeny with an estimate mu=5.14 substitutions/genome, an MRCA = 1997, and significant test of temporal signal. Model testing with a null temporal signal (*i.e.,* all sampling dates equal) phylogeny indicates temporal signal model (model=carc) was significant. We found a strong correlation between the sampling dates and the root-to-tip distances in addition to the significant temporal signal (P-value<0.05). We selected for 10000 Markov Chain Monte Carlo simulations using a ‘carc’ model to obtain the dated phylogeny.

**Supplemental Fig. 2.**
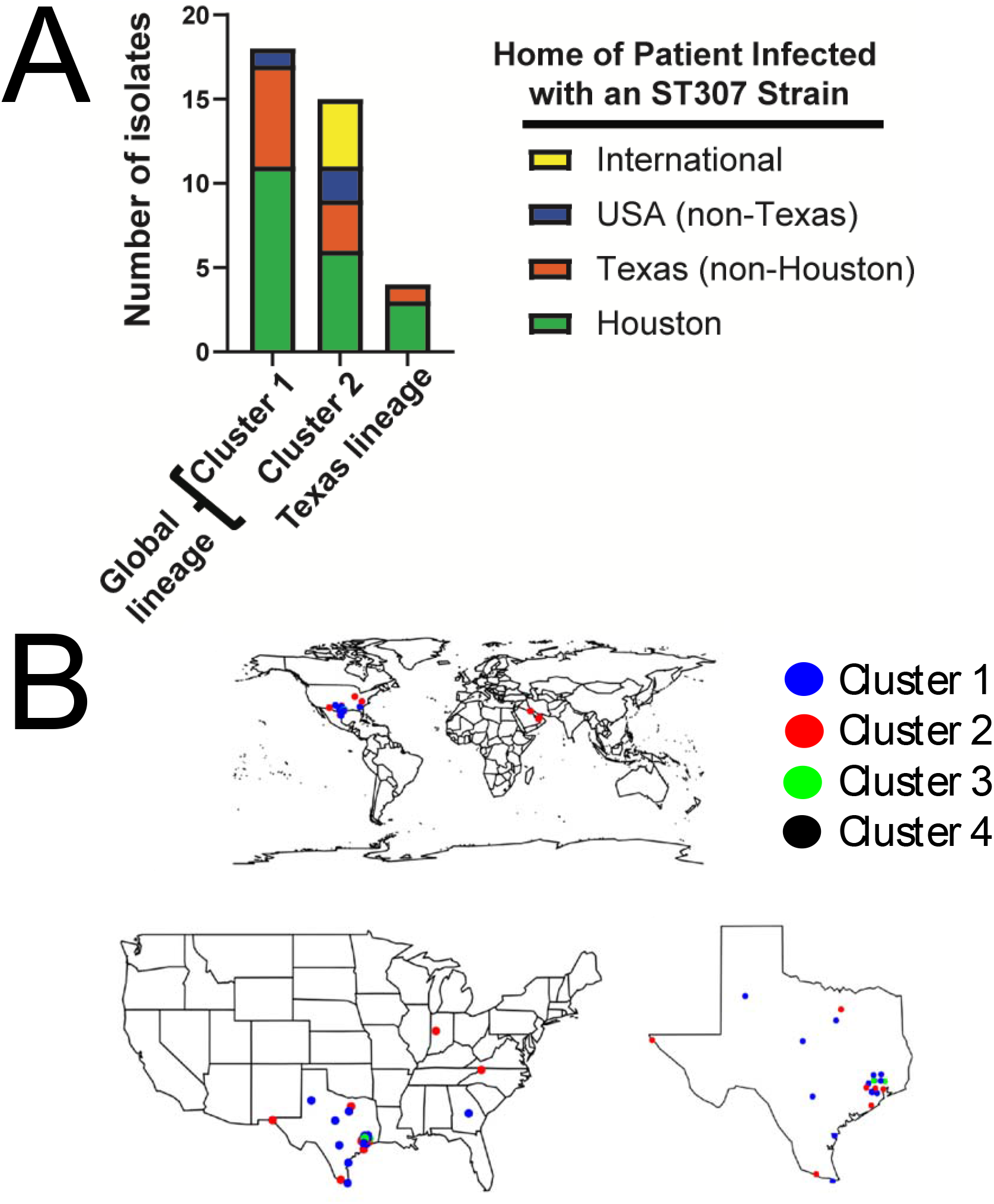
Geographic location of patient origin in which CG307 strains caused bacteremia at our institution. (A) X-axis stratifies isolates by clusters identified using hierarchical clustering. Colors of patient origin as shown in legend. (B) Geographical distribution from a world, country, and state perspective. Isolates are colored by cluster as indicated in legend.

**Supplemental Fig. 3.**
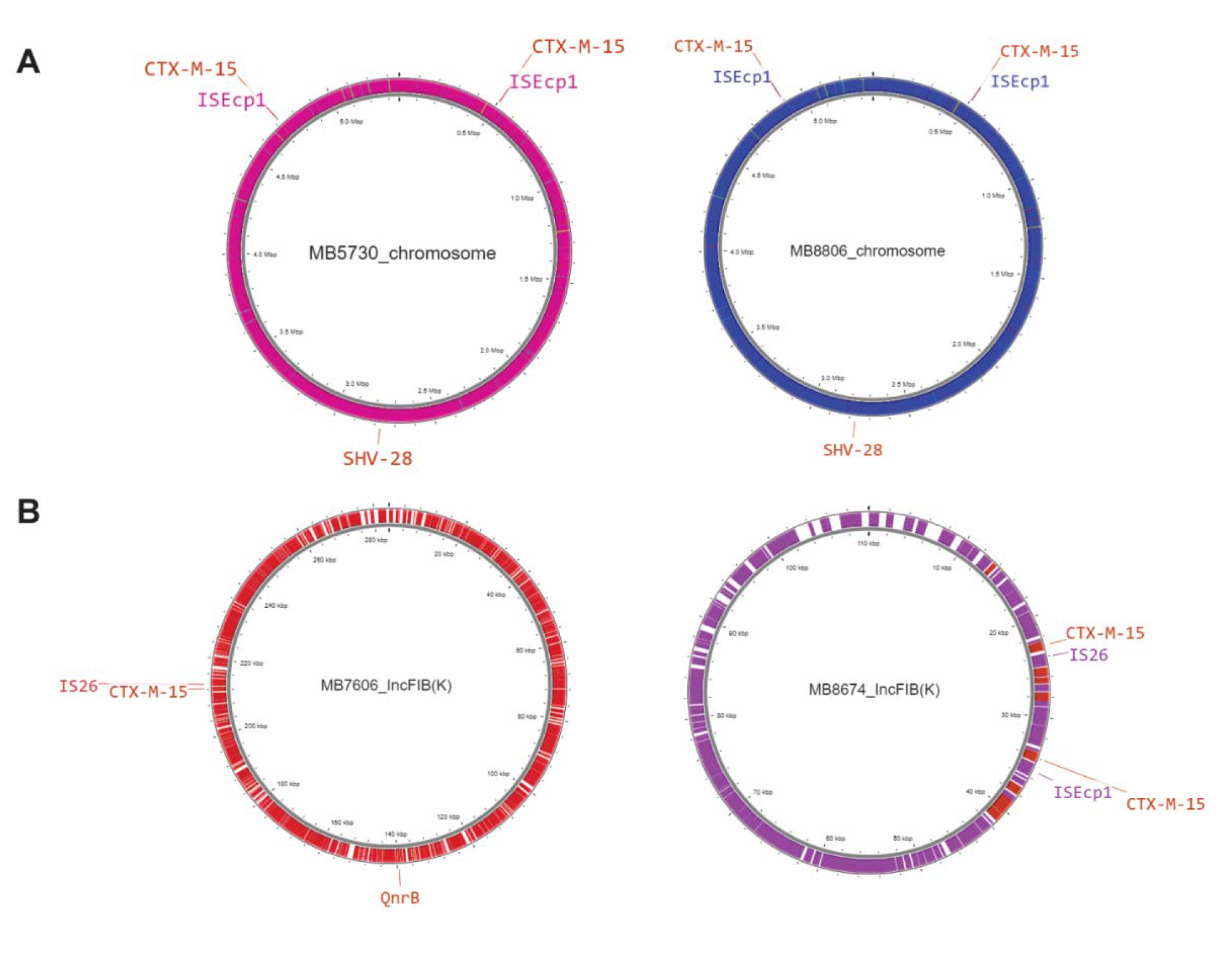
Analysis of CG307 bla_CTX-M-15_ copy number and genomic context from MDA 3GC-R*Kp* isolates. (A) CG307 Texas-specific clade isolates (*i.e.,* cluster 3 and cluster 4) that have two copies of *bla*_CTX-M-15_ present on the chromosome in association with IS*Ecp1* as previously described [49]. (B) CG307 global clade isolates (*i.e.,* cluster 1 and cluster 2) that have *bla*_CTX-M-15_ present in one or more copies on F-type multireplicon plasmids.

## Data Availability

All data produced in the present study are available upon reasonable request to the authors

## ACKNOWLEDGEMENTS

We would like to thank the MDACC clinical microbiology lab for all their hard work in identifying, handling, and transferring these pathogenic strains of interest to us for our research projects. Core grant CA016672(ATGC) and NIH 1S10OD024977-01 grant provide funding for the Advanced Technology Genomics Core (ATGC) sequencing facility at MD Anderson Cancer Center. SSA was supported by Dell Family Fund for the School of Health Professional scholarship and CTW is supported by Peter and Cynthia Hu scholarship. WCS is supported through the National Institute of Allergy and Infectious Diseases (NIAID) T32 AI141349 Training Program in Antimicrobial Resistance. Support for this study was also provided by the NIAID R21AI151536 and P01AI152999 for S.A.S. The authors acknowledge the support of the High-Performance Computing for research facility at the University of Texas MD Anderson Cancer Center for providing computational resources that have contributed to the research results reported in this paper.

